# An Analysis of Territorial Patterns in COVID-19 Mortality in France, Spain, Italy and the UK

**DOI:** 10.1101/2020.07.27.20162677

**Authors:** Roberto Zavatta

## Abstract

This paper provides an overview of territorial patterns of COVID-19 deaths in four European countries severely affected by the pandemic, Spain, France, Italy, and the United Kingdom. The analysis focuses on cumulated COVID-19 mortality at the sub-regional level, following the territorial subdivision of countries adopted by the European Union. The paper builds upon a dataset with highly granular information on COVID- 19 deaths assembled from various sources. The analysis shows remarkable differences in territorial patterns of COVID-19 mortality, both within and across the four countries reviewed. Results somewhat differ depending on the aspect considered (concentration of deaths or mortality rates) but, in general, Italy, France and Spain display significant territorial disparities, with selected sub-regions being disproportionately affected by the pandemic. Instead, the picture is more uniform in the UK, with comparatively lower differences across the various sub- regions. These findings suggest that analyses of COVID-19 mortality at the national level (and, sometimes, even at the regional level) may conceal major differences and therefore be of limited use, both analytically and from an operational viewpoint.

## 1 Introduction

Both media sources and scholarly publications analyzing the death toll caused by the COVID-19 pandemic typically focus on mortality in ‘high level’ geographical areas, i.e. at the country and/or regional (or state) levels. However, the pandemic has had a highly heterogenous spatial effect and analyses at an aggregated level may not be able to fully capture its differentiated territorial impact. This paper seeks to address the issue by providing a descriptive analysis of COVID-19 deaths at a finer geographical level.

This paper provides an assessment of territorial patterns of COVID-19 deaths in four European countries severely affected by the pandemic, Spain, France, Italy, and the United Kingdom (UK). The analysis focuses on mortality at the **sub-regional level**, i.e. for the so called NUTS3 areas as defined by the European Union (EU). This is made possible by the collection and systematization of highly granular information on COVID- 19 deaths, which was retrieved from various sources.

The paper is structured as follows: (i) Section 2 elaborates on the territorial scope of the analysis; (ii) Section 3 deals with the measurement of COVID-19 mortality and related data sources; (iii) Section 4 shows the geographical concentration of COVID-related deaths; (iv) Section 5 does the same with reference to mortality rates; and (v) Section 6 offers some concluding remarks.

## 2 Territorial Scope of the Analysis

In the 1970s, the EU established a classification of territorial entities commonly known as ‘NUTS’, which is the acronym of the French expression *Nomenclature des unités territoriales statistiques*. The classification system includes three hierarchical levels, i.e. NUTS1, corresponding to ‘macro-regions’ (e.g. North Western Italy or Occitanie), NUTS2, corresponding to regions or states (e.g. Piedmont or Bretagne), and NUTS3, corresponding to sub-regional entities. The NUTS classification tends to reflect the administrative subdivisions established within EU Member States, although in some cases a certain NUTS level may be the result of the aggregation/subdivision of administrative units. The NUTS classification is periodically revised. In this paper, reference is made to the classification adopted in 2016.^[1]^

In the four countries analyzed in this paper, the NUTS3 have different origins and characteristics. In **Spain**, NUTS3 correspond to the *provincias*, originally established by the administrative reform of 1833 (although some of them can trace their origins to much older territorial entities, such as Navarra). Spain’s NUTS3 come in different size and shape but they are generally larger than their counterparts in the other countries: the 52 Spanish *provincias* have an average area of nearly 10,000 square kilometers and a population of about 900,000 (with a median of some 600,000 inhabitants). The NUTS3 in **France** (*départements*) and **Italy** (*province*) also have a long history (in France, dating back to the end of the XVIII century), although in Italy over the last couple of decades there have been several re-organizations. NUTS3 in the two countries are similar in number (101 *départements* and 107 *province*) and population (570,000 in Italy and 660,000 in France), although French NUTS3 are more than twice the size of Italy’s (more than 6,000 vs. less than 3,000 square kilometers). In the **UK**, NUTS3 are a mix of historical counties (such as Dorset or Devon) and groupings of unitary authorities (in England and Wales) or council/district council areas (in Scotland and Northern Ireland). UK’s NUTS3 are much more numerous (179) and significantly smaller (on average, 1,400 square kilometers and 370,000 inhabitants) than in the three other countries.

**Exhibit 1.**
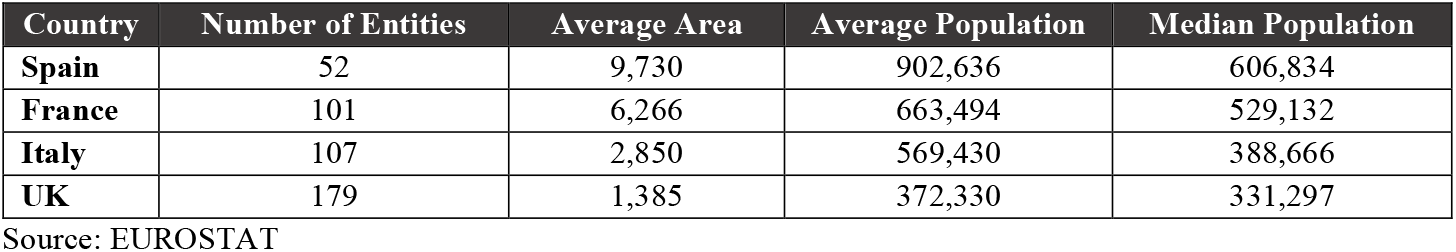
Basic Features of NUTS3.

| Country | Number of Entities | Average Area | Average Population | Median Population |
| --- | --- | --- | --- | --- |
| **Spain** | 52 | 9,730 | 902,636 | 606,834 |
| **France** | 101 | 6,266 | 663,494 | 529,132 |
| **Italy** | 107 | 2,850 | 569,430 | 388,666 |
| **UK** | 179 | 1,385 | 372,330 | 331,297 |

Source: EUROSTAT

As it will be seen below, the different size of NUTS3 entities in the four countries sometimes does have an impact on the analysis of territorial patterns of COVID-19 mortality.

### Overseas Territories

French and Spanish NUTS3 also include some overseas territories, often referred to as ‘outermost regions’. These include the five French *départments d’outre-mer* (Guadeloupe, Martinique, Guyane, La Réunion and Mayotte) as well as Spain’s Canary Islands, which are subdivided into two *provincias*. Considering that territorial contiguity is a key factor in the spreading of pandemics, these overseas NUTS3 entities have been excluded from the analysis. The same applies to Spain’s autonomous cities of Ceuta and Melilla, which, while not formally regarded as ‘outermost regions’, are located on the northern shores of the Moroccan coast and therefore are not contiguous to the rest of Spanish territory.

## 3 COVID-19 Mortality

### Measurement of COVID-19 Deaths

In the four countries covered by this paper, data on COVID-19 mortality is collected and disseminated primarily by health authorities, at the national and/or regional level. These statistics typically only refer to deaths confirmed by a molecular test (the ‘confirmed deaths’). The exclusion of deaths of individuals displaying symptoms but not tested for the virus obviously results in an underestimation of ‘real’ COVID-19 mortality. Also, in the early stages of the pandemic, data on ‘confirmed deaths’ concerned mostly or exclusively deaths occurring in hospitals. The coverage of COVID-19 mortality statistics was progressively expanded to include deaths in nursing homes and other institutions, but there are indications that gaps may have remained.^[2]^ Finally, deaths occurring at home, which typically do not involve any form of testing, are still largely unreported.^[3]^

In some countries, mortality statistics issued by public health authorities are supplemented by data resulting from population registers and death certificates. This is particularly the case of the UK, where the statistical agencies in England & Wales, Scotland and Northern Ireland also issue data on ‘certified deaths’, i.e. deaths for which there is a mention to COVID-19 in the death certificate. The statistics based on death certificates also include ‘suspect’ COVID-19 cases, and therefore are more comprehensive than the data provided by health authorities. However, there are indications that even ‘certified deaths’ may underestimate COVID-19 mortality.^[4]^

Overall, there is little doubt that currently available data underestimates the real magnitude of COVID-19 mortality. In addition, as data collection and reporting practices may somewhat differ across countries (e.g. due to differences in the coverage of deaths in nursing homes), figures for the four countries may not be fully comparable. However, this does not have a major impact on the analysis presented in this paper, as the emphasis is more on the comparison of trends rather than of absolute figures.

### Sources of Data

The analysis focuses on cumulated COVID-19 deaths occurred over the March – May 2020 period, corresponding to the peak of the pandemic in the four countries analyzed. Data on COVID-19 mortality at the NUTS3 level are not always readily available and the dataset underpinning this paper was built using various sources.^[5]^

In the case of **Spain**, the information on cumulated COVID-19 ‘confirmed deaths’ at the provincial level was primarily retrieved from the database established by Estudio Montera. In turn, this database relied on data reported by the *Comunidades Autonomas*, i.e. the NUTS2 level entities that have prime responsibility for the running of the health system, or by *Instituto de Salud Carlos III*, (ISCIII) Spain’s National Health Institute. This paper primarily refers to the situation prevailing at the end of May. However, access to Estudio Montera’s dataset also allowed to easily retrieve information on deaths at end March and end April, which in turn allowed for a diachronic assessment.

In the case of **France**, data on COVID-19 mortality in hospitals at the *département* level are published daily by *Santé publique France*, the agency in charge of epidemiological surveillance placed under the *Ministère des Solidarités et de la Santé*. This was complemented with information on deaths occurring in nursing homes and other institutions (*établissements sociaux et médico-sociaux* – ESMS), also published by *Santé publique France* but on a weekly basis and in separate publications. The information presented in this paper refers to the situation at end May.

As for the **UK**, reference is made to data on cumulated ‘certified deaths’ in the local administrative units and other similar territorial subdivisions, published by the statistical agencies in England & Wales, Scotland, and Northern Ireland. Figures refer to the cumulated deaths registered as of end of May. As the statistics on ‘certified deaths’ require more time to be compiled, the data used here may not include some deaths occurred before 31 May but not yet registered on that date. However, this is considered to have a no material effect on the analysis.

Finally, in the case of **Italy**, statistics on COVID-19 mortality at the provincial level are not systematically published by health authorities. Reference was therefore made to a study recently published by the *Istituto Superiore di Sanità* (ISS) and the National Statistical Institute (ISTAT), covering developments up to end of May. Two earlier, similar studies covered the situation up to end March and end April, which again allowed for an overtime comparison.

All in all, this paper covers nearly 143,000 COVID-19 deaths subdivided among 429 NUTS3 entities in the four countries.

**Exhibit 2.**
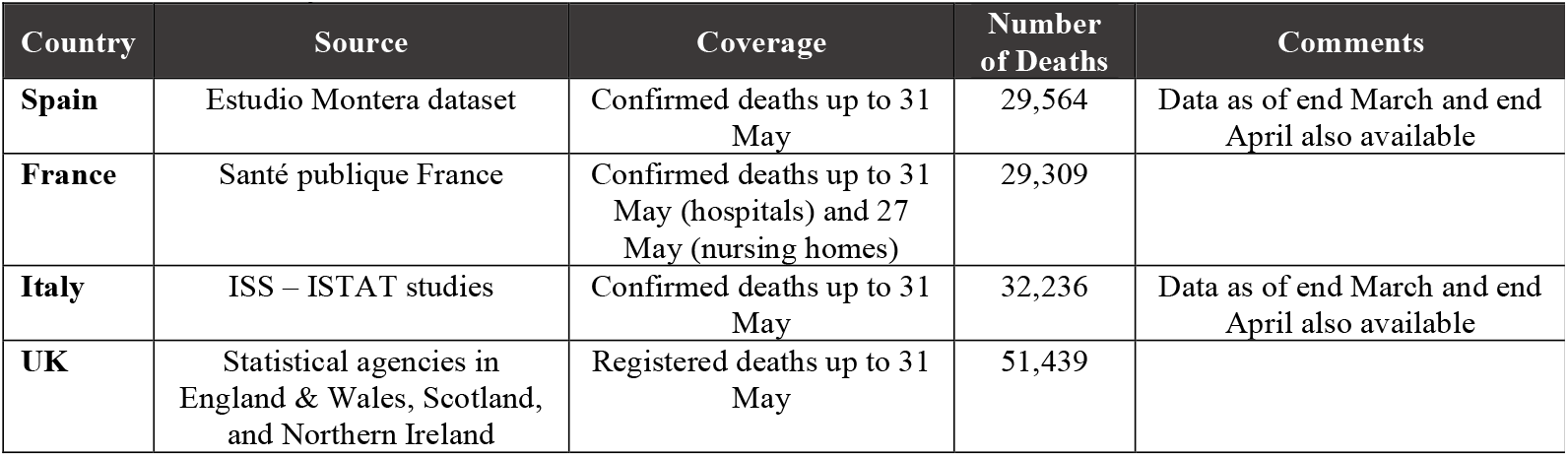
Summary of Data Utilized.

## 4 Findings – Territorial Concentration of COVID-19 Deaths

The territorial concentration of COVID-19 deaths shows significant variations across countries, with widely diverging patterns.

Concentration is fairly high in **Italy**. At the end of May, the five NUTS3 with the highest number of deaths (the ‘Top 5’), all but one located in Lombardia, accounted for more than 38% of total COVID-19 mortality nation-wide. Another 15% was accounted for by the next five highest ranking NUTS3 (the ‘Next 5’), located in Lombardia as well as in Liguria and Emilia-Romagna. All in all, the ten NUTS3 topping the ranking (the ‘Top 10’), all located in the northwestern part of the country, accounted for nearly 53% of COVID-related deaths country-wide.

Concentration is higher in **Spain**, with the Top5 NUTS3 accounting for over 60% of cumulated deaths country wide, a share increasing to 71% when the Next 5 are added. However, these figures are strongly influenced by the structural features of Spanish *provincias*, that – as mentioned above – are significantly larger than their counterparts in other countries and therefore ceteris paribus are expected to have more deaths. Indeed, Madrid and Barcelona, the two areas with the highest death toll, are quite sizeable territorial entities, cumulatively accounting for more than a quarter of total Spanish population. In comparison, Milano and Bergamo, the two Italian *province* with the highest death tolls, are much smaller, with a population of slightly more than 7% nation-wide.

Concentration is lowest in the **UK**. The Top 5 NUTS3 account for less than 9% of the COVID-19 total death toll, and the share increases to only 15% when the Next 5 areas are considered. To some extent, this is also due to the structural features of UK’s NUTS3, which are much smaller than in other countries and therefore are expected to have fewer deaths each. However, this is only part of the story. Indeed, even if we consider the top 20 NUTS3, their share in cumulated deaths remains a fairly low 26%, i.e. lower than the share of Madrid alone.

**France** is an intermediate case. The Top 5 NUTS3, including Paris and its inner metropolitan area (the so called *petite couronne*) as well as one eastern *département*, account for little more than 30% of total French COVID-19 deaths. The share of the Next 5, also mostly from the Île de France plus the NUTS3 encompassing Lyon and another eastern *département*, is close to 19%, bringing the share of cumulated COVID-19 deaths in the Top 10 close to 50%.

**Exhibit 3.**
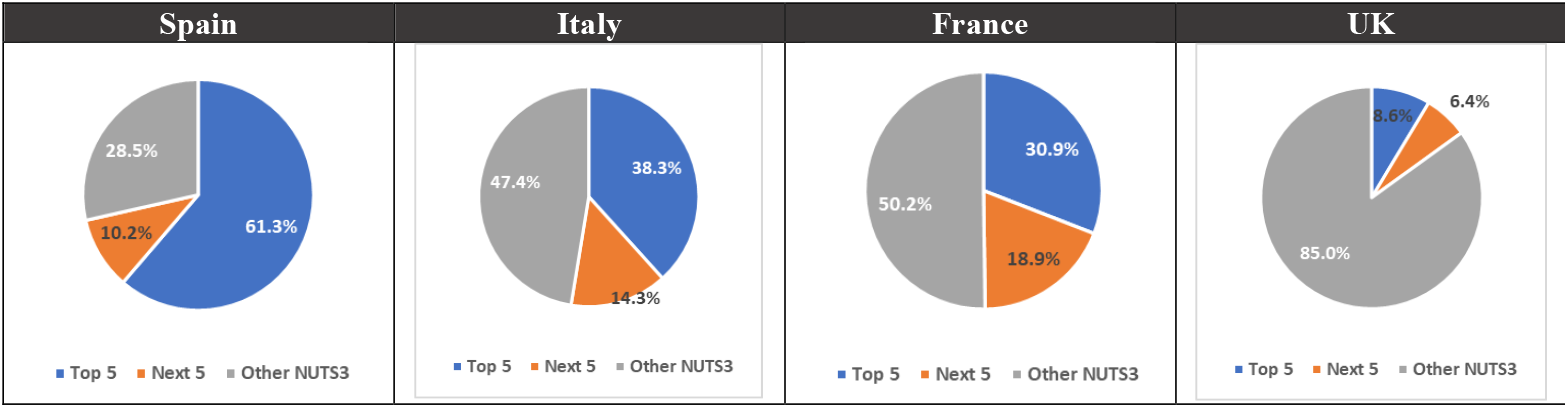
Overview of Territorial Concentration of COVID-19 Deaths.

### Box 1

**Changes in Territorial Concentration Overtime**

In the case of Spain and Italy, the availability of mortality data at various time intervals allows for an assessment of changes in concentration over time. In **Spain**, concentration was highest at the end of March, when the Top 10 *provincias* accounted for 77% of total COVID-19 deaths. The share declined to 71% by end April and stayed at the same level at end May. The list of *provincias* comprising the Top 10 remained broadly the same, with Madrid, Barcelona, Ciudad Real and Bizkaia constantly topping the list, and only few new entries (namely, Girona, ranking 12th at end March but subsequently raising to the 7th/8th position) and exists (namely, Álava, ranking 7th at end March and subsequently dropping out of the Top 10, i.e. 16th at end May).

In **Italy**, the decline in concentration was more marked. At the end of March, the Top 10 NUTS3 accounted for nearly 65% of country-wide deaths, with the top two *province*, Bergamo and Brescia, accounting alone for 29%. By end May, the Top 10 share had declined by more than ten percentage points, to about 53%. The composition of the Top 10 also shows some significant changes, with Milano overtaking Bergamo as the area with the largest number of deaths and the entry of other two large urban areas, Turin and Genova, ranking respectively fourth and eighth.

## 5 Findings – Territorial Patterns in Mortality Rates

A complementary and more accurate picture of territorial developments in COVID-related mortality can be obtained by looking at ‘mortality rates’, i.e. the ratio between the number of deaths and the population in any given geographical area. Expressed in terms of deaths per 100,000 inhabitants, mortality rates are a standardized metric, not influenced by the size of NUTS3.

Once again, there are significant variations across the four countries. In France and, especially, in Italy, mortality rates reached quite high levels in a limited number of NUTS3 and the pandemic scarcely affected many other areas. In contrast, COVID-19-related deaths are more evenly spread nationwide in Spain and especially in the UK.

In **France**, the first outbreak was in the Eastern part of the country and the pandemic quickly reached the capital and the rest of the Île de France region. There were other localized outbreaks (e.g. in Corse-du-Sud in the early days), but they were of much smaller size, and the south and south western regions were largely spared. As a result, the five most affected *départements* (the ‘Top 5’), with mortality rates ranging from 110 up to nearly 190 deaths per 100,000, are all located in the North Eastern part of the country. In these areas mortality rates are twenty to forty times bigger than those recorded in the five least affected areas (the ‘Bottom 5’), all located in South Western France, which reported fewer than 5 deaths per 100,000. The skewed territorial distribution of mortality rates is confirmed by summary statistics, with the average being nearly twice the median value (38 vs. 22 deaths per 100,000). The corresponding coefficient of variation (CV)^[6]^ is fairly high, at 98%

The situation is even more polarized in **Italy**, where mortality rates have reached very high levels in the initial Codogno cluster (in Lombardy’s Lodi province) and surrounding areas.^[7]^ Indeed, at the end of May three of the Top 5 *province* (located in Lombardy and Emilia-Romagna) recorded mortality rates in excess of 300 per 100,000, by far the highest values recorded at the NUTS3 level across the four countries. These values are more than one hundred times bigger than those found in the Bottom 5 areas (mostly in Sicily), which reported just one to three deaths per 100,000. The extremely skewed distribution of mortality rates in Italy is again captured by the difference between the average and median values, with the former being more than double than the latter (53 vs. 24 deaths per 100,000), while the CV is quite high, at 130%.

### Box 2

**Differences in Mortality Rates Within Italian Regions**

In Italy, stark differences in mortality rates are sometimes found also <u>within</u> regions, i.e. at the NUTS2 level. This is particularly the case of the **Emilia-Romagna** region, where the 336 deaths per 100,000 recorded in the northwestern province of Piacenza (which lies just across the Po river at short distance for the initial Codogno cluster) was fifteen times the rate recorded in the eastern Ravenna province, 21 deaths per 100,000. An even starker contrast is found in the **Marche** region, were the 142 deaths per 100,000 recorded in Pesaro Urbino are a multiple of the 5 deaths per 100,000 in the Ascoli Piceno province, which yet lies just 200 kilometers to the south. Even in **Lombardy**, where the vast majority of provinces were heavily affected, there are some notable differences. In particular, the 58 deaths per 100,000 recorded in Varese are about one sixth of the 280/310 deaths per 100,000 found in Cremona, Lodi and Bergamo, which are just 100 kilometers away.

In **Spain**, the pandemic also had a differentiated territorial impact, but the distribution of mortality rates is comparatively less unbalanced. In the Top 5 *provincias* (all in central Spain), mortality rates were typically in the order of 130 – 160 deaths per 100,000, with only Ciudad Real reaching the level of 221 deaths per 100,000. These figures are about ten times the values displayed by the Bottom 5 NUTS3 (mostly in Southern Spain), which recorded between 8 and 13 deaths per 100,000. Spain’s more uniform territorial distribution of mortality rates is confirmed by summary statistics, with the average value of 63 deaths per 100,000 being higher but not too dissimilar from the median value of 48, while the CV is 76%.

An even more uniform pattern is found in the **UK**. Leaving aside the two areas with the highest rates (North & West Norfolk and Brent, with respectively 181 and 143 deaths per 100,000) and half a dozen areas in Scotland and Northern Ireland scarcely affected by the pandemic, NUTS3 mortality rates display a smooth, monotonically decreasing pattern from 120 deaths per 100,000 to 30 per 100,000. The uniform distribution is symbolized by the limited difference between the average and the median values, with the latter being actually greater than the former (76 vs 74 deaths per 100,000. Consequently, the CV is a modest 37%.

**Exhibit 4.**
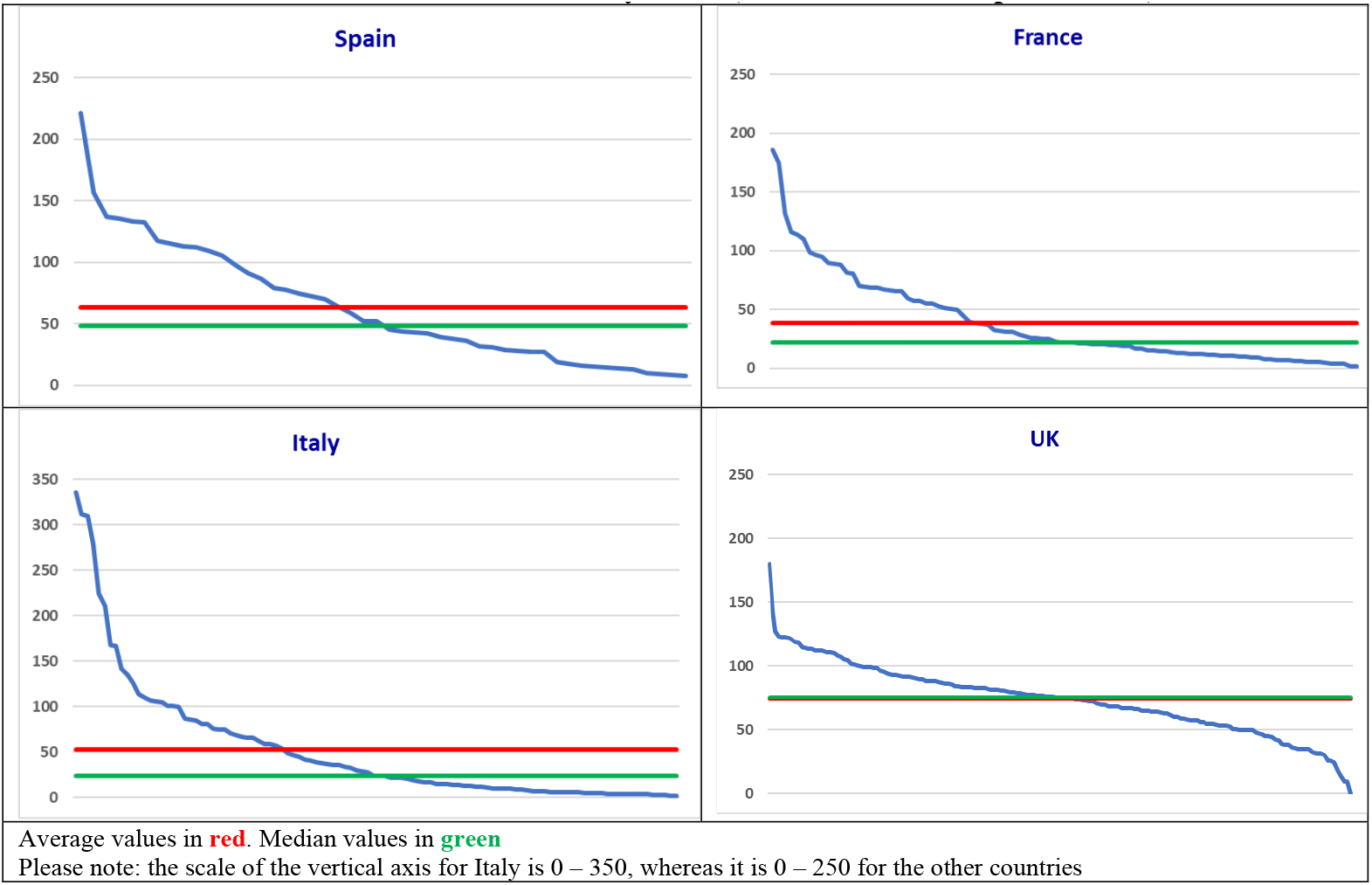
Territorial Distribution of Mortality Rates (COVID-19 Deaths per 100,000)

## 6 Concluding Remarks

The results presented above clearly show remarkable differences in territorial patterns of COVID-19 mortality, both within and across the four countries reviewed. Results somewhat differ depending on the aspect considered (concentration of deaths or mortality rates) but, in general, Italy, France and Spain display significant territorial disparities. Instead, the picture is comparatively more uniform in the UK. These findings confirm the initial intuition of the paper, i.e. that analyses of COVID-19 mortality at the national level (and, sometimes, even at the regional level) may conceal major differences and therefore be of limited use, both analytically and from an operational viewpoint.

The reasons for these territorial differences remain to be investigated. Prima facie, the most heavily affected NUTS3 areas appear to be those where the initial outbreaks took place (e.g. Lodi, Haut-Rhin), suggesting a declining rate of propagation, also because of the lockdown measures adopted. However, the impact of other, more structural aspects (such as population density or the share of older population) must also be considered. The availability of the highly granular information on COVID-19 deaths assembled for this paper will greatly facilitate a more comprehensive analysis, making it possible to match mortality data with a range of socio- economic indicators available at the NUTS3 level.

## Data Availability

Should it be of any interest, the Author would be delighted to provide the full dataset underpinning the paper, so that other researchers may use the data for further research on the topic.

## SUPPLEMENTARY MATERIALS

## ANNEX A – DETAILED RESULTS

**Exhibit A.1.**
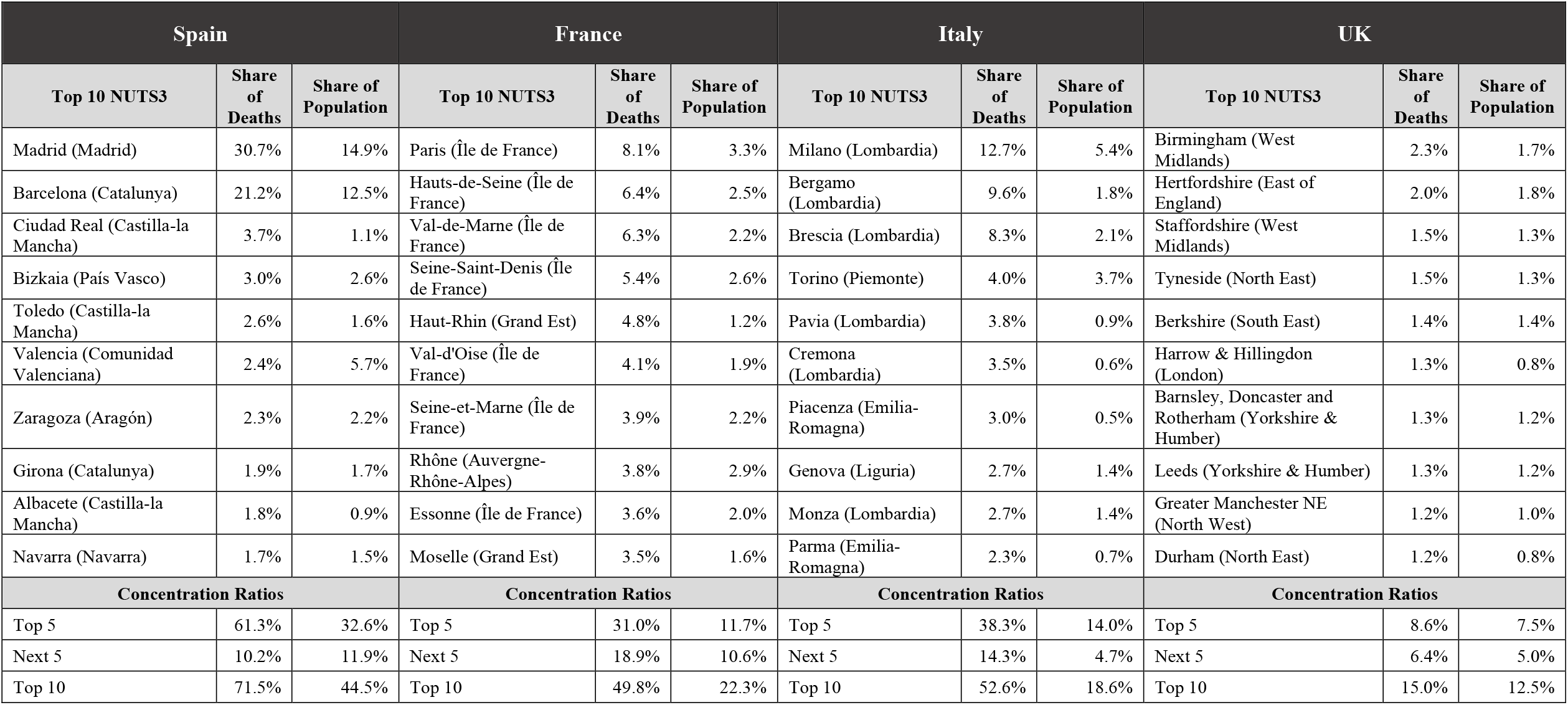
Territorial Concentration of COVID-19 Deaths.

| Spain |  |  | France |  |  | Italy |  |  | UK |  |  |
| --- | --- | --- | --- | --- | --- | --- | --- | --- | --- | --- | --- |
| Top 10 NUTS3 | Share of Deaths | Share of Population | Top 10 NUTS3 | Share of Deaths | Share of Population | Top 10 NUTS3 | Share of Deaths | Share of Population | Top 10 NUTS3 | Share of Deaths | Share of Population |
| Madrid (Madrid) | 30.7% | 14.9% | Paris (Île de France) | 8.1% | 3.3% | Milano (Lombardia) | 12.7% | 5.4% | Birmingham (West Midlands) | 2.3% | 1.7% |
| Barcelona (Catalunya) | 21.2% | 12.5% | Hauts-de-Seine (Île de France) | 6.4% | 2.5% | Bergamo (Lombardia) | 9.6% | 1.8% | Hertfordshire (East of England) | 2.0% | 1.8% |
| Ciudad Real (Castilla-la Mancha) | 3.7% | 1.1% | Val-de-Marne (Île de France) | 6.3% | 2.2% | Brescia (Lombardia) | 8.3% | 2.1% | Staffordshire (West Midlands) | 1.5% | 1.3% |
| Bizkaia (País Vasco) | 3.0% | 2.6% | Seine-Saint-Denis (Île de France) | 5.4% | 2.6% | Torino (Piemonte) | 4.0% | 3.7% | Tyneside (North East) | 1.5% | 1.3% |
| Toledo (Castilla-la Mancha) | 2.6% | 1.6% | Haut-Rhin (Grand Est) | 4.8% | 1.2% | Pavia (Lombardia) | 3.8% | 0.9% | Berkshire (South East) | 1.4% | 1.4% |
| Valencia (Comunidad Valenciana) | 2.4% | 5.7% | Val-d'Oise (Île de France) | 4.1% | 1.9% | Cremona (Lombardia) | 3.5% | 0.6% | Harrow & Hillingdon (London) | 1.3% | 0.8% |
| Zaragoza (Aragón) | 2.3% | 2.2% | Seine-et-Marne (Île de France) | 3.9% | 2.2% | Piacenza (Emilia-Romagna) | 3.0% | 0.5% | Barnsley, Doncaster and Rotherham (Yorkshire & Humber) | 1.3% | 1.2% |
| Girona (Catalunya) | 1.9% | 1.7% | Rhône (Auvergne-Rhône-Alpes) | 3.8% | 2.9% | Genova (Liguria) | 2.7% | 1.4% | Leeds (Yorkshire & Humber) | 1.3% | 1.2% |
| Albacete (Castilla-la Mancha) | 1.8% | 0.9% | Essonne (Île de France) | 3.6% | 2.0% | Monza (Lombardia) | 2.7% | 1.4% | Greater Manchester NE (North West) | 1.2% | 1.0% |
| Navarra (Navarra) | 1.7% | 1.5% | Moselle (Grand Est) | 3.5% | 1.6% | Parma (Emilia-Romagna) | 2.3% | 0.7% | Durham (North East) | 1.2% | 0.8% |
| Concentration Ratios |  |  | Concentration Ratios |  |  | Concentration Ratios |  |  | Concentration Ratios |  |  |
| Top 5 | 61.3% | 32.6% | Top 5 | 31.0% | 11.7% | Top 5 | 38.3% | 14.0% | Top 5 | 8.6% | 7.5% |
| Next 5 | 10.2% | 11.9% | Next 5 | 18.9% | 10.6% | Next 5 | 14.3% | 4.7% | Next 5 | 6.4% | 5.0% |
| Top 10 | 71.5% | 44.5% | Top 10 | 49.8% | 22.3% | Top 10 | 52.6% | 18.6% | Top 10 | 15.0% | 12.5% |

**Exhibit A.2.** Territorial Distribution of Mortality Rates (COVID-19 Deaths per 100,000)

| France |  | Italy |  | Spain |  | UK |  |
| --- | --- | --- | --- | --- | --- | --- | --- |
| Top 5 NUTS3 | Rates | Top 5 NUTS3 | Rates | Top 5 NUTS3 | Rates | Top 5 NUTS3 | Rates |
| Haut-Rhin (Grand Est) | 185.8 | Piacenza (Emilia-Romagna) | 335.7 | Ciudad Real (Castilla-la Mancha) | 221.4 | North & West Norfolk (East of England) | 180.6 |
| Territoire de Belfort (Bourgogne-Franche-Comté) | 174.4 | Cremona (Lombardia) | 311.5 | Cuenca (Castilla-la Mancha) | 156.8 | Brent (London) | 143.2 |
| Val-de-Marne (Île de France) | 131.5 | Lodi (Lombardia) | 309.3 | Madrid | 136.6 | Walsall (West Midlands) | 127.6 |
| Hauts-de-Seine (Île de France) | 116.0 | Bergamo (Lombardia) | 279.0 | Albacete (Castilla-la Mancha) | 135.5 | Harrow & Hillingdon (London) | 123.2 |
| Vosges (Grand Est) | 114.0 | Pavia (Lombardia) | 224.4 | Soria (Castilla y León) | 133.0 | Wirral (North West) | 123.1 |
| Bottom 5 NUTS3 | Rates | Bottom 5 NUTS3 | Rates | Bottom 5 NUTS3 | Rates | Bottom 5 NUTS3 | Rates |
| Dordogne (Nouvelle-Aquitaine) | 4.1 | L'Aquila (Abruzzo) | 3.0 | Cádiz (Andalucía) | 12.6 | Derry City and Strabane (Northern Ireland) | 17.9 |
| Landes (Nouvelle-Aquitaine) | 3.9 | Oristano (Sardegna) | 2.5 | Murcia (Murcia) | 9.8 | Fermanagh and Omagh (Northern Ireland) | 13.7 |
| Lot-et-Garonne (Nouvelle-Aquitaine) | 3.6 | Ragusa (Sicilia) | 2.2 | Huelva (Andalucía) | 9.2 | Inverness & Nairn and Moray, Badenoch & Strathspey (Scotland) | 9.3 |
| Ariège (Occitanie) | 1.3 | Trapani (Sicilia) | 1.6 | Lugo (Galicia) | 8.2 | Orkney Islands (Scotland) | 9.1 |
| Lozère (Occitanie) | 1.3 | Palermo (Sicilia) | 1.4 | Almería (Andalucía) | 7.6 | Western Isles (Scotland) | 0.0 |
| Summary Statistics | Rates | Summary Statistics | Rates | Summary Statistics | Rates | Summary Statistics | Rates |
| Average | 38.2 | Average | 46.1 | Average | 63.2 | Average | 74.0 |
| Median | 21.8 | Median | 20.0 | Median | 48.2 | Median | 75.6 |
| Coefficient of Variation | 98% | Coefficient of Variation | 137% | Coefficient of Variation | 76% | Coefficient of Variation | 37% |

**Exhibit A.3.**
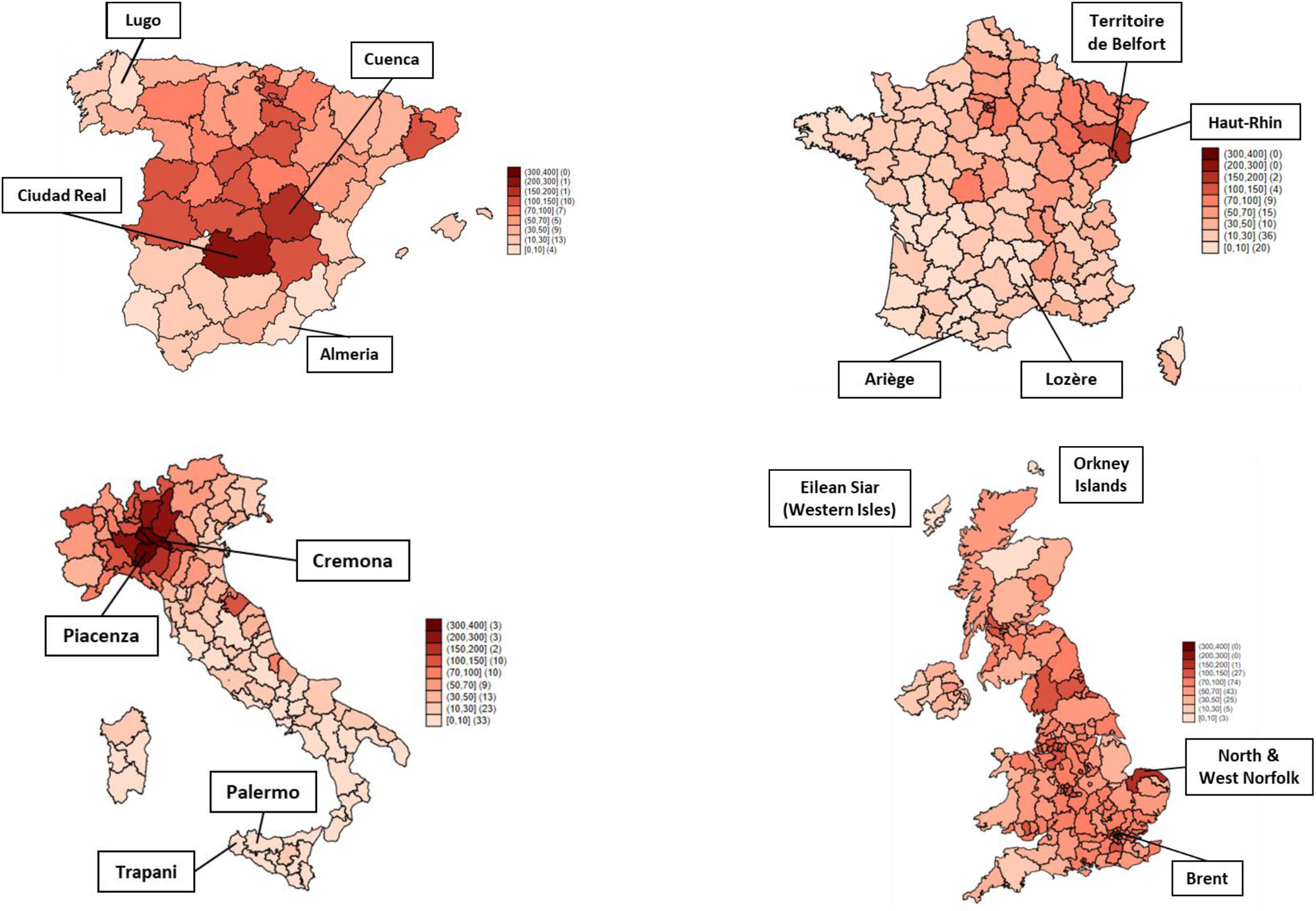
Territorial Distribution of Mortality Rates (COVID-19 Deaths per 100,000)

## ANNEX B – DATA SOURCES

### B.1 Spain

Data on COVID-19 cumulated deaths used in this paper are mostly taken from the dataset built by Estudio Montera (hereinafter, the ‘Estudio Montera Dataset’), complemented with own estimates for the three provincias in the Pais Vasco.

The Estudio Montera Dataset is accessible at https://github.com/montera34/escovid19data. The database includes information on deaths as well as a host of other relevant variables (number of cases, tests performed, etc.). For each variable, the dataset also shows the original sources of data, typically health authorities in the various *Comunidades Autonomas* and the ISCIII. The data collection and compilation work performed by Estudio Montera is extremely valuable as information on COVID-19 deaths at the provincial level is sometimes not easily accessible. Therefore, Estudio Montera’s contribution is once again gratefully acknowledged.

The analysis presented here relies on cumulated deaths data at end March, end April and end May.^[8]^ Regarding the latter, in some cases available data do not refers to the situation as of 31 May but to earlier dates. In the case of the A Coruña, province (Asturias), data refers to 23 May. Considering that this province has relatively low levels of COVID-19 mortality and the bulk of deaths occurred in April, the underestimation of mortality entailed by the use of older data is minimal, probably in the order of few units.

The situation is different for the three *provincias* in the País Vasco (Álava, Bizkaia and Gipuzkoa), as the most recent figures included in the Estudio Montera Dataset are significantly older, referring to 13 May. Therefore, the end of May figures used for this paper were estimated by applying the proportion of deaths in each province on 13 May resulting from the Estudio Montera Dataset to the total number of COVID-19 deaths reported by regional authorities in their daily epidemiological report.^[9]^

Overall, the total number of COVID-19 deaths considered for the analysis at the end of May was 29,564, whereas the deaths at end March and end April were, respectively, 9,176 and 25,776.

### B.2 France

The data on COVID-19 deaths used for this paper comes from two sources, namely: (i) the statistics on COVID-19 deaths occurring in hospitals; and (ii) the statistics on deaths occurring in the ESMS, which nursing homes (*établissements d’hébergement pour personnes âgées dépendantes* - EHPAD) and other institutions.

Data on deaths in hospitals are collected by *Santé publique France* and made available through an online database updated daily (https://www.coronavirus-statistiques.com/stats-globale/coronavirus-nombre-de-morts-par-departement/). The database was last accessed on 7 June 2020 and at the date of 31 May it showed a total of 18,413 deaths (excluding 51 deaths in the overseas *départements* not covered by this paper).

Data on COVID-19 deaths in ESMS are also collected by *Santé publique France* and published in a series of regional epidemiological reports. These reports are published on a weekly basis and they are available through a dedicated section of *Santé publique France*’s website.^[10]^ The website was last accessed on 29 May and the regional reports available at that date covered the situation up to the week ending on 27 May.

The format of regional reports shows some variations and details on the deaths in each *département* are not always readily available. Accordingly, in some cases the relevant information had to be estimated. In particular:

- In the case of three regions, Corsica, Hauts-de-France and Nouvelle-Aquitaine, the epidemiological reports reviewed only provided the total number of deaths in ESMS. The figures at the *département* level were estimated by applying the proportion of deaths in hospitals derived from the online database mentioned above;
- The report on the Île de France region does provides figures at the *département* level but they also include the deaths of people living in ESMS that occurred in hospitals, therefore partly overlapping with data provided in the online database. Since deaths in hospitals were reported to account for 22% of total ESMS deaths region-wide, figures for individual *départements* were computed by discounting this proportion.

Based on the above, COVID-19 deaths in ESMS were estimated at 10,896 as of 27 May. When added to the deaths in hospitals, this brings the total number of deaths considered in the analysis at 29,309.

### B.3 Italy

In Italy, data on COVID-19 deaths at the provincial level are not systematically published. Information is provided by some regions (e.g. Toscana) and some figures are from time to time published in the media, but there is no way to obtain a timely and comprehensive picture of mortality at the NUTS3 level.^[11]^

Therefore, this paper primarily relies on the information provided in a study published in early July by ISS and ISTAT and covering developments up to the end of May.^[12]^ The purpose of the study is to assess the impact of the COVID-19 pandemic on overall mortality, by comparing COVID-19 deaths at the provincial level with total mortality over the same period as well as with the mortality recorded in previous years. The study provides the provincial breakdown of 32,236 COVID-19 deaths occurred as of 31 May. This figure is smaller than the total number of COVID-19 deaths recorded over the same period in Italy (32,981). However, the difference is quite small (745 deaths, i.e. about 2%) and it is not deemed to affect the validity of the analysis.^[13]^

This paper could also rely on previous, similar studies also published by ISS and ISTAT, which covered developments in mortality up 31 March^[14]^ and up to 30 April.^[15]^ The existence of studies with comparable information at three points in time allowed to assess the evolution of territorial patterns in COVID-19 mortality.

A final note concerns Sardinian provinces. In 2016, a reform reduced the number of *province* from eight to five and also involved the transfer of some territory among previous and new administrative units. The reform is not yet reflected in EU statistics on NUTS3, which are still based on the previous territorial architecture. As population data for the computation of mortality rates per 100,000 was taken from EU statistics, there could be small discrepancies due to (minor) changes in provincial boundaries. For the same reason, the Sud Sardegna province is lumped together with the *Città Metropolitana di Cagliari*. In practice, this means that this paper considered 106 NUTS3 entities instead of 107. As Sardinia was one of the least affected areas by COVID-19, none of these adjustments had any appreciable effect on the analysis.

### B.4 United Kingdom

As anticipated in the text, in the case of the UK this paper relied on data on ‘certified deaths’ collected and published by statistical agencies. In particular:

- In the case of England and Wales reference was made to data published by the Office for National Statistics (ONS) and concerning the deaths registered up to 29 May, subdivided by local authority;^[16]^
- For Scotland, reference was made to the dataset published by the National Records of Scotland (NRC) and concerning deaths registered up to 31 May, subdivided by council area of usual residence;^[17]^
- Regarding Northern Ireland, data comes from the Northern Ireland Statistics and Research Agency (NISRA) and concern the deaths registered in the week up to 29 May, subdivided by local government district.^[18]^

The preference for ‘certified deaths’ statistics is obviously due to their wider coverage compared with data on ‘confirmed deaths’ issued by health authorities. Since ‘certified deaths’ statistics require more time for their compilation, information is available with some delay. Therefore, UK data may not include some deaths occurred before end May but not yet registered on that date. Overall, the analysis presented in this paper considers a total on 51,439 deaths.

## Competing interests

The author declares no competing interests nor financial relationships with any organizations that might have an interest in the submitted work. The work for the paper was entirely self-financed by Economisti Associati, a fully independent economic and public policy consultancy.

## Acknowledgments

Data on COVID-19 mortality in Spain was taken from a pre-existing dataset developed by Estudio Montera. The collaboration of Pablo Rey and his colleagues is gratefully acknowledged. Silvia Beghelli and Costanza Fileccia assisted in the collation and/or review of data for France and Italy. Carlotta Moiso did the same for the United Kingdom and prepared the maps in supplementary materials.

## Notes

### Competing Interest Statement

The authors have declared no competing interest.

### Funding Statement

No external funding has been received for the submitted work.

### Author Declarations

There are no details to be provided.

